# PD-1^high^CXCR5^–^CD4^+^ Peripheral Helper T (Tph) cells Promote Tissue-Homing Plasmablasts in COVID-19

**DOI:** 10.1101/2021.03.13.21253527

**Authors:** Hiromitsu Asashima, Subhasis Mohanty, Michela Comi, William E. Ruff, Kenneth B. Hoehn, Patrick Wong, Inessa Cohen, Sarah Coffey, Khadir Raddassi, Omkar Chaudhary, Avraham Unterman, Brinda Emu, Steven H. Kleinstein, Ruth R. Montgomery, Akiko Iwasaki, Charles S. Dela Cruz, Naftali Kaminski, Albert C. Shaw, David A. Hafler, Tomokazu S. Sumida

**Affiliations:** Department of Neurology, Yale School of Medicine, New Haven, CT, USA; Department of Immunobiology, Yale School of Medicine, New Haven, CT, USA; Section of Infectious Diseases, Department of Internal Medicine, Yale School of Medicine, Yale University, New Haven, CT, USA; Department of Pathology, Yale School of Medicine, New Haven, CT, USA; Section of Pulmonary, Critical Care and Sleep Medicine Section, Department of Internal Medicine, School of Medicine, Yale University, New Haven, CT, USA; Inter-Departmental Program in Computational Biology and Bioinformatics, Yale University, New Haven, CT, USA; Department of Internal Medicine, Yale School of Medicine, New Haven, CT, USA; Howard Hughes Medical Institute, Chevy Chase, MD, USA

**Keywords:** PD-1^high^CXCR5^–^CD4+ peripheral helper T (Tph) cells, tissue-homing plasmablasts, IFNγ, COVID-19

## Abstract

A dysregulated immune response against coronavirus-2 (SARS-CoV-2) plays a critical role in the outcome of patients with coronavirus disease 2019 (COVID-19). A significant increase in circulating plasmablasts is characteristic of COVID-19 though the underlying mechanisms and its prognostic implications are not known. Here, we demonstrate that in the acute phase of COVID-19, activated PD-1^high^CXCR5^−^CD4^+^ T cells, peripheral helper T cells, (Tph) are significantly increased and promote inflammatory tissue-homing plasmablasts in patients with stable but not severe COVID-19. Analysis of scRNA-seq data revealed that plasmablasts in stable patients express higher levels of tissue-homing receptors including *CXCR3*. The increased Tph cells exhibited “B cell help” signatures similar to that of circulating T follicular helper (cTfh) cells and promoted B cell differentiation *in vitro*. Compared with cTfh cells, Tph cells produced more IFNγ, inducing tissue-homing chemokine receptors on plasmablasts. Finally, expansion of activated Tph cells was correlated with the frequency of CXCR3^+^ plasmablasts in the acute phase of patients with stable disease. Our results demonstrate a novel role for Tph cells in acute viral immunity by inducing ectopic, antibody secreting plasmablasts.

## Introduction

SARS-CoV-2 causes a wide spectrum of symptoms ranging from asymptomatic infections to acute respiratory distress syndrome (ARDS)^1,2^. COVID-19 is the clinical manifestation of SARS-CoV-2 infection and it has become clear that a dysregulated immune response against SARS-CoV-2 is central in determining disease severity. Increased frequencies of circulating neutrophils with low frequencies of natural killer cells or CD4^+^ T cells are characteristic of peripheral blood immune profiles observed in patients with COVID-19^3–6^. The generation of a robust antibody response is critical for clearing the virus and alterations in B cell lineages with the expansion of plasmablasts are reported both in the blood and bronchoalveolar lavage of patients with COVID-19^6–8^. The increase in plasmablasts, which can represent up to 30% of circulating B cells, is observed in a subset of patients comparable to acute Ebola or dengue virus infections^9,10^.

During the acute phase of COVID-19, activated B cells differentiate into plasmablasts that migrate to target inflammatory tissues, especially the lungs^11^, indicating that trafficking patterns of plasmablasts are important in the disease course. These plasmablasts express tissue-specific chemokines together with adhesion molecules and produce organ-specific protective antibodies for viral control^12,13^. This rapid antibody response is critical as the early containment of virus reduces the risk of cytokine storm syndrome with excessive accumulation of immune cells in the lung parenchyma with ARDS. Despite this important role, the control of expansion and migration capacity of plasmablasts in patients with COVID-19 remains poorly understood.

During the adaptive immune response, T follicular helper (Tfh) cells play crucial roles in B cell differentiation^14,15^. Circulating CXCR5^+^CD4^+^ T cells, termed cTfh cells, support B cell differentiation in blood while Tfh cells located in lymphoid tissues allow B cell differentiation in lymph nodes^16^. Intriguingly, while the plasmablast response correlates with the cTfh responses in subjects recovered from COVID-19^17^, there is no correlation between the frequency of cTfh cells and that of plasmablasts in symptomatic patients with COVID-19^6^. Moreover, in the acute phase of SARS-CoV-2 infection, a striking absence of germinal centers in lymphoid organs was reported, similar to SARS, but activation-induced cytidine deaminase (AID)-expressing B cells are still preserved^18^. These data suggest that activated helper T cells other than Tfh cells support the differentiation of B cells at extra-follicular regions in acute phase of COVID-19.

To examine the mechanism of T cell regulation of plasmablast differentiation in COVID-19, we investigated the characteristics of B and T cells in patients by single cell RNA-seq (scRNA-seq) and flow cytometry datasets. Here, we report that PD-1^high^CXCR5^−^CD4^+^ T cells, so called peripheral helper T (Tph) cells^19,20^, are significantly increased and positively correlated with the frequency of plasmablasts in peripheral blood in patients with COVID-19. These Tph cells exhibit “B cell help” signatures to a similar degree as cTfh cells, but also express more inflammatory chemokine receptors including *CCR2* and *CCR5. In vitro* experiments indicate that PD-1^high^CXCR5^−^ Tph cells have higher IFNγ production, which promotes CXCR3 expression and differentiation of plasmablasts. Finally, we demonstrate that CXCR3^+^ tissue-homing plasmablasts are significantly increased in patients with stable COVID-19 while they are decreased in patients with severe disease. These findings provide a mechanism for the increase of plasmablasts apart from Tfh cells in the acute phase of COVID-19. Thus, the induction of plasmablast trafficking to inflammatory sites by activated PD-1^high^CXCR5^−^ Tph cells is important for disease control in the acute phase of COVID-19.

## Results

### Expanded plasmablasts express higher tissue-homing molecules in patients with stable COVID-19

We first confirmed that the frequency of plasmablasts is significantly increased in the peripheral blood in patients with COVID-19 by flow cytometry^3,21^ (Figure 1a, Supplemental Table 1, 2). To further understand the characteristics of B cells in patients with COVID-19, we next analyzed previously collected scRNA-seq data^5^ and sub-clustered B cells (total 13550 cells from 31 samples) into eight clusters according to gene expression (Figure 1b, 1c, Supplemental Figure 1a, 1b, Supplemental Table 3). We identified naïve B cells (*MS4A1*^+^*IGHD*^+^), germinal center-like B cells (*MS4A1*^+^*NEIL1*^+^; GC-like B cells), intermediate memory B cells (*IGHD*^+^*CD27*^+^), memory B cells (*MS4A1*^+^*CD27*^+^), and two plasma cells clusters: plasmablasts (*MZB1*^+^*CD38*^+^) and Ki67^+^ plasmablasts (*MZB1*^+^*CD38*^+^*MKI67*^+^), in accordance with a previous report^22^ (plasmablasts; 912 cells and Ki67^+^ plasmablasts; 537 cells). GC-like B cells also express *CD9*^23,24^ (Supplemental Figure 1b). Additionally, we were able to identify two additional clusters, namely, FCRL5^+^ B cells (*MS4A1*^+^*FCRL5*^+^) and CD1c^+^ B cells (*MS4A1*^+^*CD1C*^+^). FCRL5^+^ B cells also express higher levels of *ITGAX* and *ZEB2* than the other B cells, and this cluster was similar with atypical B cells or double-negative (DN) cells^25–27^ (Supplemental Figure 1b). CD1c^+^ B cells express higher levels of *CD52* compared to the other B cell subclusters, which implies that gene expression signatures of this cluster resemble marginal zone-like B cells^28,29^. There was no significant difference in the clonal diversity or the frequency of unmutated clones between the two clusters of plasmablasts (Supplemental Figure 1c, 1d). Both plasmablast clusters expressed IgG, which indicated that they had undergone class-switching (Figure 1d). Ki67^+^ plasmablasts had a significantly lower frequency of somatic hypermutations (SHM) in COVID-19 subjects compared with the other plasmablasts (Figure 1e). The proportion of GC-like B cells was significantly decreased and FCRL5^+^ B cells were increased in patients with progressive COVID-19. These observations imply that the extrafollicular response is related to B cell activation in patients with COVID-19^30^ (Supplemental Figure 1e). To further examine the characteristics of each single cell RNA cluster, we determined gene expression levels as related to B cell function^31^ (Supplemental Figure 1f). It was of interest that the expression level of *CXCR3*, the receptor necessary for trafficking to sites of inflammation, was upregulated in plasmablast clusters, especially in Ki67^+^ plasmablasts. Homing receptors^32^, which promote homing to lymph nodes (*CCR7, SELL*), were highly expressed in healthy control plasmablasts, while Ki67^+^ plasmablasts in stable COVID-19 patients expressed tissue-homing molecules (*CXCR3, CCR2*) (Figure 1f, 1g). Taken together, Ki67^+^ plasmablasts trafficking to inflammatory sites are a unique population expanded in stable patients in contrast to patients with severe COVID-19, and these acute plasmablast responses might be beneficial for hosts to prevent viral expansion at inflammatory tissues.

**Figure 1.**
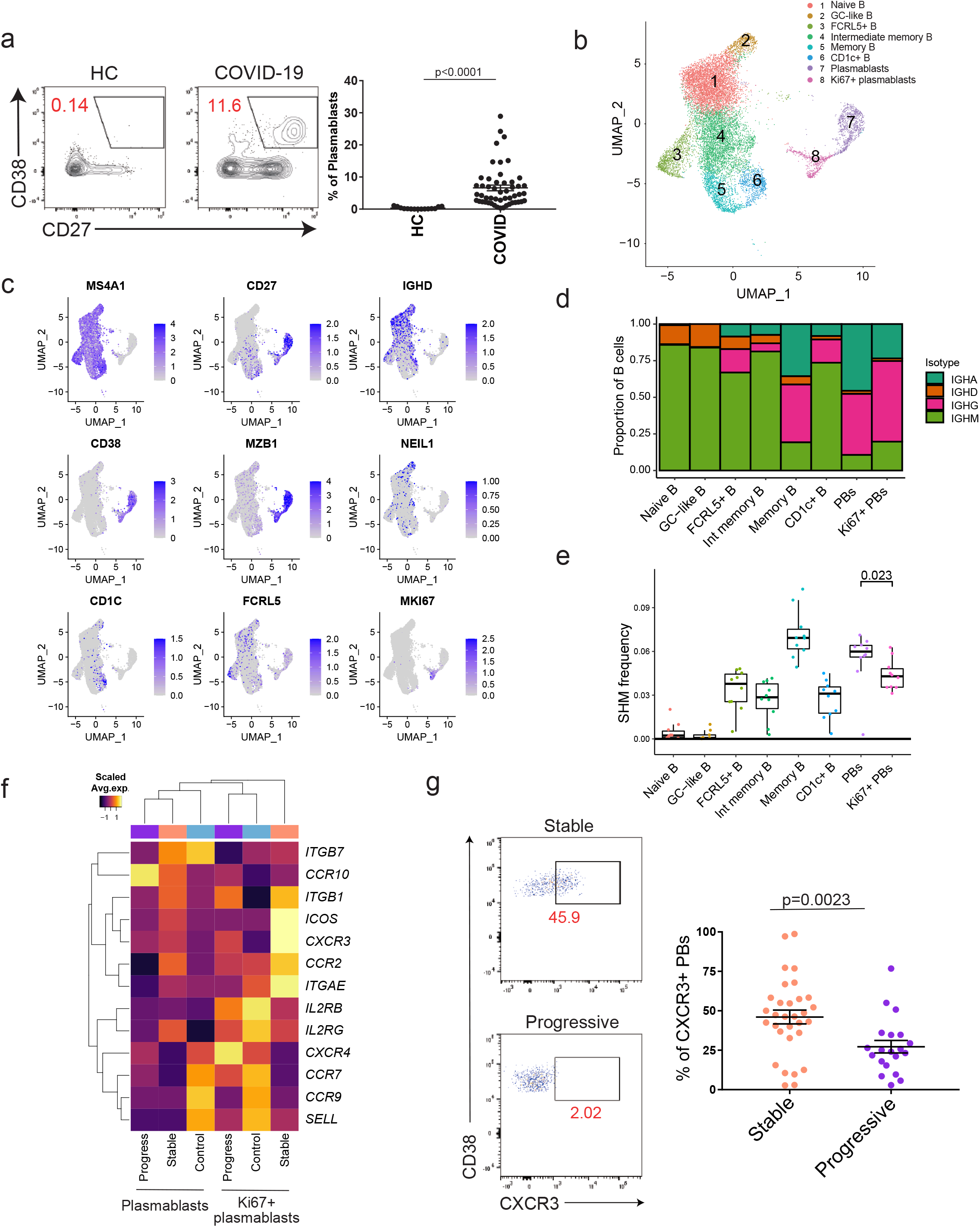
The divergent immunological features of B cells in stable and progressive COVID-19 patients. Plasmablasts of patients with stable COVID-19 express inflammatory tissue-homing receptors. **a**, Representative flow data of CD19^+^CD27^+^CD38^+^ plasmablasts (left). The proportion of plasmablasts between healthcare workers (HCs) (n=15) and both stable and progressive COVID-19 patients (COVID) (n=51) were evaluated by two-tailed unpaired Student’ s t-test were evaluated (right). **b**, UMAP representation of sub-clustered B cells from HCs (n = 13) and COVID-19 samples (n = 18 from 10 patients). Eight subclusters were identified. **c**, Canonical cell markers for cluster delineation. Data are colored according to expression levels. **d**, Fractional abundance of IGHA (dark green), IGHD (orange), IGHG (pink), and IGHM (light green) cells in each cluster. PBs denote plasmablasts. **e**, Frequency of somatic hypermutation (SHM) in each cluster. Each dot denotes a patient (combined early and late samples, n=10). A Wilcoxon test was evaluated, and p value is reported above plasmablast clusters. PBs denote plasmablasts. **f**, Heatmap of tissue-homing receptors^32^ among HC, stable COVID-19 (stable), and progressive COVID-19 (progressive) in clusters of both plasmablasts and Ki67+ plasmablasts. Average expression per subject for each gene is shown. **g**, Representative flow data of CXCR3 expression on CD19^+^CD27^+^CD38^+^ plasmablasts in COVID-19 patients (left). The proportions of CXCR3^+^ plasmablasts between stable (n= 31) and progressive (n=20) COVID-19 patients were evaluated by two-tailed unpaired Student’ s t-test (right).

### PD-1^high^CXCR5^−^ Tph cells are significantly increased in patients with COVID-19 and have a unique gene expression profile

Tfh cells provide essential B cell help during the adaptive immune response, and circulating Tfh (cTfh) cells share functional properties with Tfh cells^16^. Notably, PD-1^high^CXCR5^−^ Tph cells, that are reported to be associated with extra follicular B cell differentiation^20^, were significantly increased in the blood of COVID-19 patients, in addition to significant increase in PD1^high/int^CXCR5^+^ Tfh cells (Figure 2a, Supplemental Figure 2a, 2b). Tph cells were barely detected in healthy blood and moreover, correlated in COVID-19 patients with the frequency of circulating plasmablasts (Figure 2b, Supplemental Figure 2c) but not with clinical characteristics (age, body mass index (BMI), and sex) (Supplemental Figure 3a-c). The increase of PD-1^high^CXCR5^−^ Tph cells and the positive correlation between these cells and plasmablasts in the acute phase of COVID-19 patients were validated from another dataset^6^ (Supplemental Figure 4a-c).

**Figure 2.**
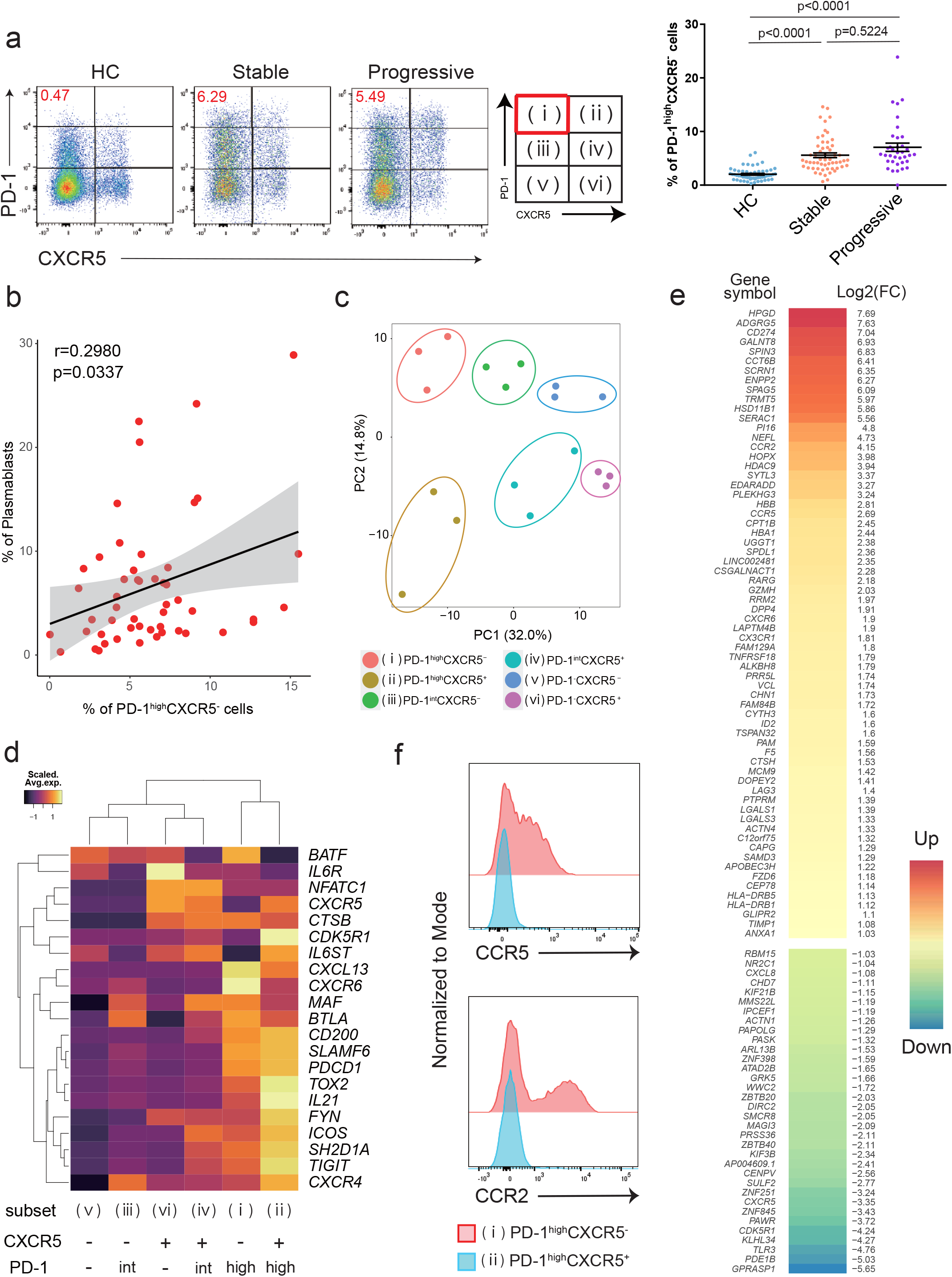
The characteristics of PD-1^high^CXCR5^−^ Tph cells. PD-1^high^CXCR5^−^ Tph cells have characteristic gene expressions and are increased in COVID-19 patients. **a**, Representative flow data of PD-1^high^CXCR5^−^ Tph cells in each group (left), the proportion of these T cells among healthcare workers (HC) (n=55), stable COVID-19 patients (Stable) (n=56), and progressive patients (Progressive) (n=36). One-way ANOVA with Dunn’ s multiple comparisons tests were performed to evaluate differences (right). **b**, Correlation between PD-1^high^CXCR5^−^ Tph cells (percentage of CD3^+^CD4^+^CD45RA^−^ memory T cells) and plasmablasts (percentage of CD19^+^ B cells) in COVID-19 patients (both stable and progressive, n=51). Linear regression is shown with 95% confidence interval (gray area). Correlation statistics is two-tailed Spearman’ s rank correlation test. **c**, Principal component analysis (PCA) of RNA-seq transcriptomes (n=3, COVID-19 patients). Based on the expression levels of PD-1 and CXCR5, six subsets (i)-(vi) were evaluated **d**, Heatmap of Tfh-related genes19 among six subsets of memory CD4+ T cells. **e**, Clustered heatmap of 100 genes that were differentially expressed (left column) in PD-1^high^CXCR5^−^ Tph cells compared with cTfh cells (PD-1^high^CXCR5^+^ Tph cells) (|Log2FC| >1, FDR <0.05). The right column shows the log2 fold change for PD-1^high^CXCR5^−^ Tph cells compared to cTfh cells. **f**, Representative flow data of CCR5 and CCR2 expression on PD-1^high^CXCR5^−^ Tph cells compared with PD-1^high^CXCR5^+^ Tfh cells.

To further characterize these T helper populations, six subsets of memory CD4^+^ T cells (CD45RA^−^ CD4^+^ T cells), categorized by PD-1 and CXCR5 expression levels, were sorted and gene expression profiles were examined by bulk RNA-seq (Figure 2c). Principal component analysis (PCA) placed PD-1^high^CXCR5^−^ Tph cells at a distinctive position relative to other subsets, indicating a unique gene expression profile. PD-1^high^CXCR5^−^ Tph cells had similar expression of Tfh-related genes^19^ compared to PD-1^high^CXCR5^+^ Tfh cells and expressed molecules including *MAF, TIGIT*, SLAMF6, and *IL21*, which are important for Tfh functions (Figure 2d). PD-1^int^CXCR5^+^ Tfh cells also expressed these genes, but had less *ICOS* expression than PD-1^high^CXCR5^+^ Tfh cell, suggesting that PD-1^high^CXCR5^+^ Tfh cells are more activated cTfh cells^14^. We identified 100 genes that were differentially expressed in PD-1^high^CXCR5^−^ Tph cells compared with PD-1^high^CXCR5^+^ Tfh cells (|Log2FC| >1, FDR <0.05) (Figure 2e); PD-1^high^CXCR5^−^ Tph cells showed marked upregulation of tissue-resident chemokine receptors, including *CCR2, CCR5* and *CX3CR1* (Figure 2f, Supplemental Figure 3d). Additionally, we surveyed the expression of T cell lineage genes, and found that Th1-like signatures (*CXCR3, TBX21, STAT1*) were highly expressed in PD-1^high^CXCR5^−^ Tph cells (Supplemental Figure 3e). Thus, PD-1^high^CXCR5^−^ Tph cells exhibit “B cell help” signatures to a similar degree as cTfh cells, but with a unique gene expression profile.

### PD-1^high^CXCR5^−^ Tph cells promote B cell differentiation, and produce more IFNγ than PD-1^high^CXCR5^+^ Tfh cells

We examined whether PD-1^high^CXCR5^−^ Tph cells promote B cell differentiation *in vitro*. As with PD-1^high^CXCR5^+^ Tfh cells, these cells induce memory B cells into plasmablasts, and initiated IgG production (Figure 3a, 3b). Stimulation with anti-CD3/CD28 antibodies induced greater IFNγ and IL-10 production from PD-1^high^CXCR5^−^ Tph cells (Figure 3c), which is of interest as IFNγ is known to upregulate *CXCR3* expression during B cell differentiation^33^. We observed that *CXCR3* expression on plasmablasts was upregulated in a dose dependent manner by IFNγ while IL-10 was not (Figure 3d, 3e). Moreover, besides *CXCR3*, other inflammatory tissue-homing receptors such as *CCR2* were upregulated, but *CXCR4* was not (Figure 3f). These data indicate that PD-1^high^CXCR5^−^ Tph cells promote B cell differentiation and have a capacity to induce plasmablasts which express homing-molecules for inflammatory tissues via IFNγ production.

**Figure 3.**
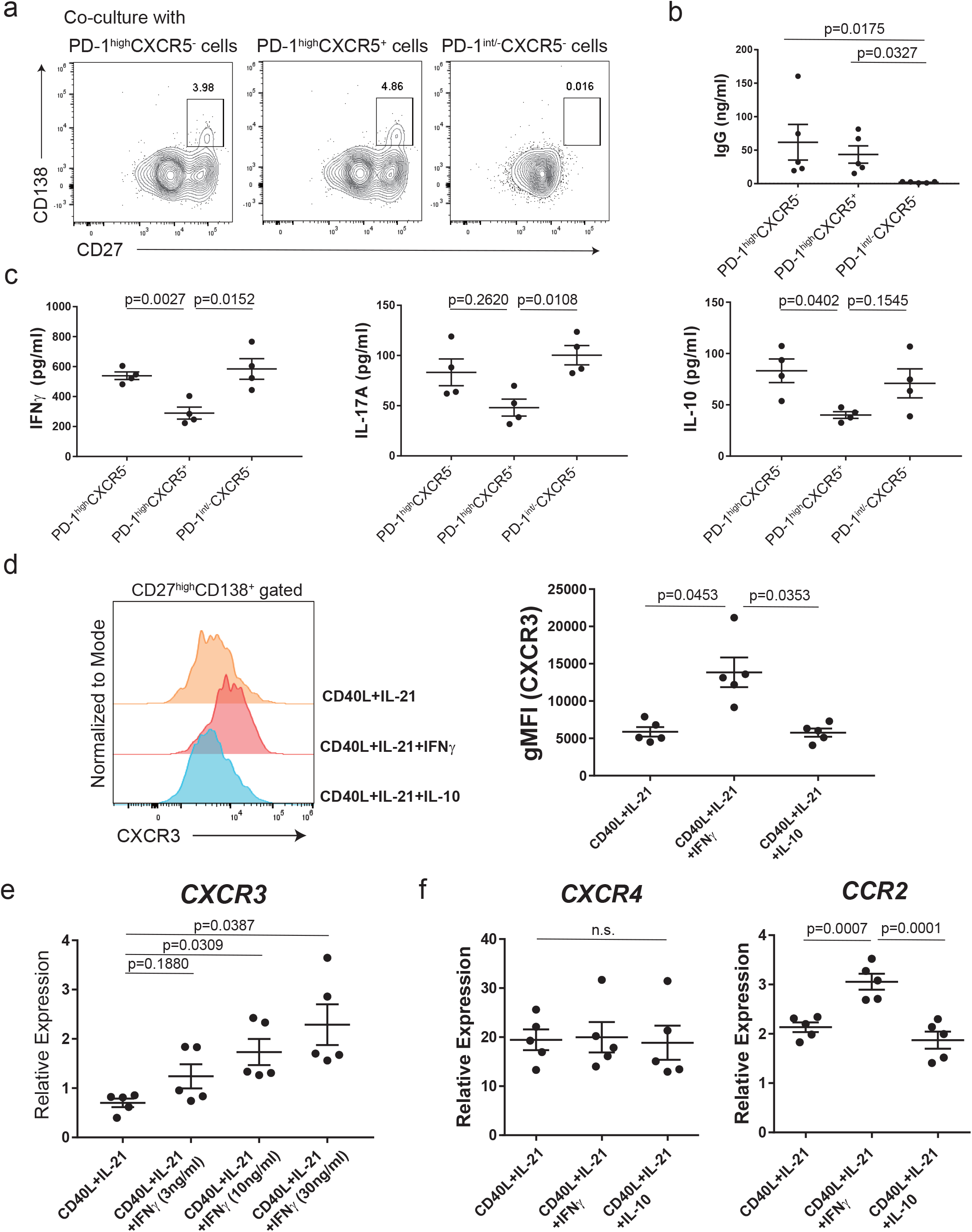
The functions of PD-1^high^CXCR5^−^ Tph cells. PD-1^high^CXCR5^−^ Tph cells promote B cell differentiation and produce IFNγ much higher than cTfh cells, which affects the expression levels of tissue-homing receptors on plasmablasts. **a-b**, Each T cell subset and autologous CD20^+^CD27^+^ B cells were sorted and co-cultured with SEB and LPS for 7 days (n=5, COVID-19 patients). Representative flow data of CD27^high^CD138^+^ plasma cells after co-culture (**a**). IgG concentrations in supernatants of co-cultures were evaluated by One-way ANOVA with Dunn’ s multiple comparisons tests (**b**). **c**, Sorted T cells (n=4, COVID-19 patients) were stimulated with anti-CD3/28 (each 1 μg/ml) for 48 hrs, then cytokine production levels were measured (IFNγ, IL-17A, IL-10). **d-f**, Sorted CD20^+^CD27^+^ memory B cells (n=5, healthy donors) were cultured with CD40L (0.05 ng/ml), IL-21 (20ng/ml), and IL-10 (10 ng/ml) or different concentration of IFNγ (0, 3, 10, 30 ng/ml) for 7 days (n=5, healthy controls). Representative histogram of flow data for CXCR3 expression on plasma cells (**d**, left), CXCR3 gMFI was evaluated by One-way ANOVA with Tukey’ s multiple comparisons tests (**d**, right). After 7 days in culture, CD19^+^CD27 ^+^CD138^+^ plasma cells were sorted and gene expression measured relative to B2M by qPCR (**e, f**). The expression levels were evaluated by One-way ANOVA with Tukey’ s multiple comparisons tests (**c-f**).

### The proportion of activated PD-1^high^CXCR5^−^ Tph cells are significantly increased in stable as compared to severely ill patients with COVID-19 and are positively correlated with CXCR3^+^ plasmablasts

To investigate the characteristics of PD-1^high^CXCR5^−^ Tph cells *in vivo*, we evaluated the specific gene signatures of this subset (Supplemental Table 4). Compared to the other five subsets in memory CD4^+^ T cells, *CXCR6, LAG3*, and *PRR5L* were significantly upregulated, while *CHD7, ZBTB20, ZNF251, GRK25*, and *GPRASP1* were significantly downregulated in PD-1^high^CXCR5^−^ Tph cells (Figure 4a, 4b). Flow cytometry analysis also showed the same trend of upregulation of LAG3 and CXCR6 expressions (Supplemental Figure 5a, 5b). Embedding these gene lists with CD4^+^ T cell clusters from our scRNA-seq data set^5^, we found that dividing CD4^+^ T cells best fit with these signatures (Figure 4c). We previously reported that dividing CD4^+^ T cells share the characteristics of HLA-DR^+^CD38^+^ activated T cells^5^, and indeed, more than half of the activated CD4^+^ T cells were in the PD-1^high^CXCR5^−^ Tph cell population (Figure 4d). In contrast, 15% of PD-1^high^CXCR5^−^ Tph cells exhibited an activated state among all COVID-19 samples (data not shown), with a significantly higher proportion in patients with stable disease as compared to patients with progressive disease (Figure 4e). We confirmed that these observations were not highly confounded by known risk factors for disease severity, such as age, BMI, and sex (Supplemental Figure 5c-e).

Finally, we observed a positive correlation between activated PD-1^high^CXCR5^−^ Tph cells and CXCR3^+^ plasmablasts (Figure 4f). There was no difference in the proportion of PD-1^high^CXCR5^−^ Tph cells between stable and progressive patients at baseline, but interestingly they increased in progressive patient over time and reached significance at two weeks after the baseline (Supplemental Figure 5f, Supplemental Table 5). In contrast, the significantly elevated frequency of activated PD-1^high^CXCR5^−^ Tph cells in stable patients at baseline became less significant over time (Supplemental Figure 5g). These data demonstrate that prompt increased HLA-DR^+^CD38^+^ activated PD-1^high^CXCR5^−^ Tph cells are one of the early responses in stable patients, which is also positively correlated with tissue-homing CXCR3^+^ plasmablasts.

**Figure 4.**
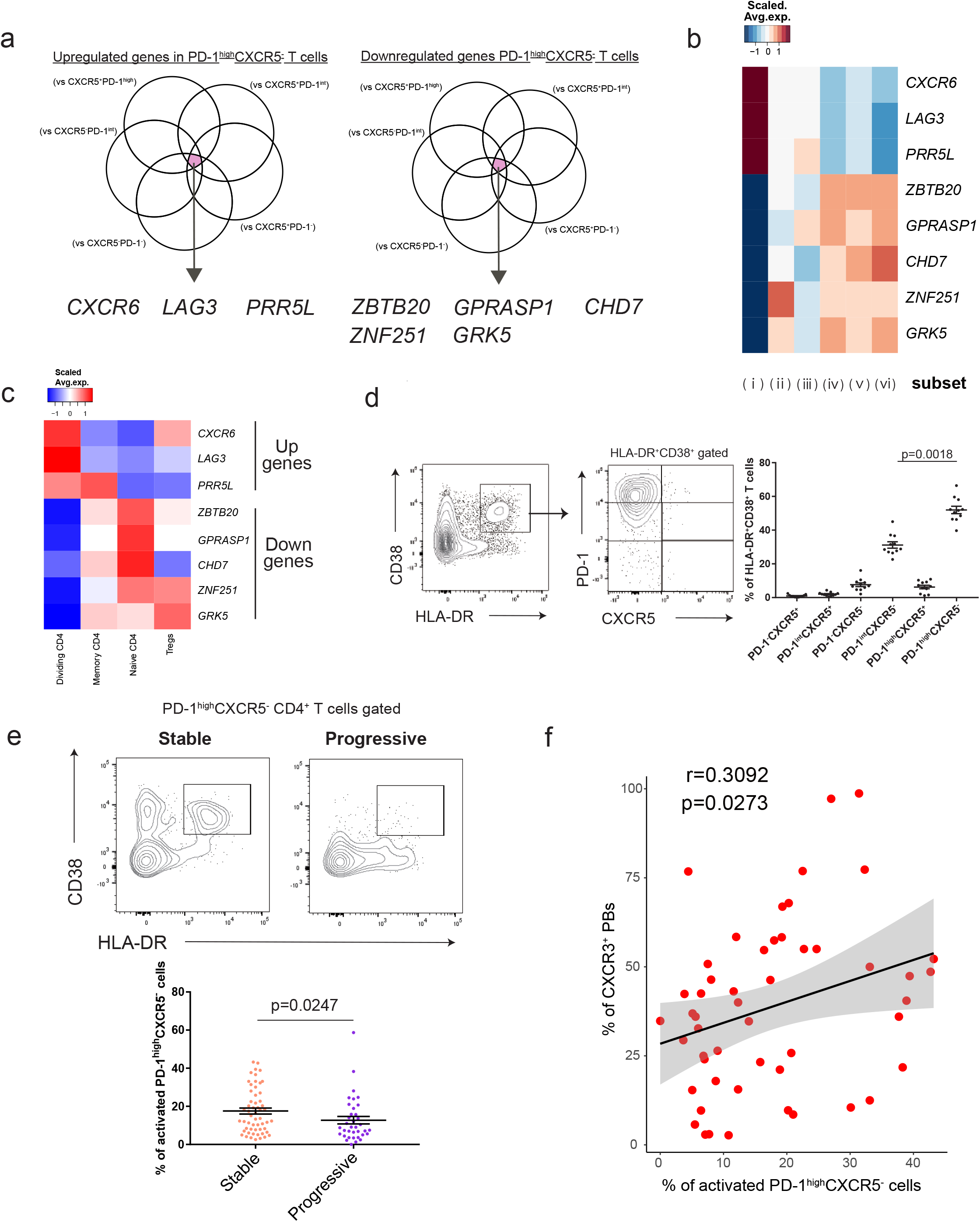
Activated PD-1^high^CXCR5^−^ Tph cells are significantly increased in stable COVID-19 patients and positively correlated with CXCR3^+^ plasmablasts. **a**, Venn diagrams showing the overlapped genes among those significantly upregulated (Log2FC>1, FDR<0.05) (left) and downregulated (Log2FC<-1, FDR<0.05) (right) in PD-1^high^CXCR5^−^ Tph cells compared with five subsets as indicated. **b**, Heatmap of PD-1^high^CXCR5^−^ Tph cells-related genes (selected in a) among each T cell subset from RNA-seq data. **c**, Heatmap of PD-1^high^CXCR5^−^ Tph cells-related genes (selected in a) among each T cell cluster of our scRNA-seq dataset reported^5^. **d**, Representative flow data for each T cell subset among HLA-DR^+^CD38^+^CD45RA^−^CD4^+^ T cells (left), and their proportions were evaluated by One-way ANOVA with Dunn’ s multiple comparisons tests (right). COVID-19 samples which have more than 5% of HLA-DR^+^CD38^+^ T cells among memory CD4^+^ T cells were evaluated (n=11). **e**, Representative flow data of HLA-DR^+^CD38^+^ activated cells in PD-1^high^CXCR5^−^ Tph cells between stable and progressive COVID-19 patients (up). The proportions of activated PD-1^high^CXCR5^−^ Tph cells were evaluated (stable; n= 56, progressive; n= 36) by two-tailed unpaired Student’ s t-test (down). **f**, Correlation between activated PD-1^high^CXCR5^−^ Tph cells (percentage of CD3^+^CD4^+^CD45RA^−^ memory T cells) and CXCR3^+^ plasmablasts (percentage of CD19^+^CD27^+^CD38^+^ plasmablasts) (both stable and progressive, n=51). Linear regression is shown with 95% confidence interval (gray area). Correlation statistics is two-tailed Spearman’ s rank correlation test.

## Discussion

Here, we performed a bidirectional-analyses between T cells and B cells in patients with COVID-19 by both bulk and scRNA-seq studies with B-cell receptor (BCR) repertoire analysis and *in vitro* assays. PD-1^high^CXCR5^−^ Tph cells were increased among the PBMCs of patients with COVID-19 and were positively correlated with the frequency of plasmablasts. These PD-1^high^CXCR5^−^ Tph cells had a similar gene profile with cTfh cells focused on “B cell help” signatures and promoted B cell differentiation *in vitro*. Moreover, CXCR3^+^ tissue-homing plasmablasts were markedly increased in patients with stable but not severely ill patients with COVID-19 where we hypothesize that activated PD-1^high^CXCR5^−^ Tph cells promoted these plasmablasts via IFNγ production. Thus, these data suggest that tissue homing plasmablasts are important for the clinical outcome of patients with COVID-19 (Supplemental Figure 6).

PD-1^high^CXCR5^−^ Tph cells were first described in the synovial fluid of rheumatoid arthritis and the peripheral blood of patients with systemic lupus erythematosus (SLE)^19,34^, indicating an important role in autoimmune diseases with tissue specific, ectopic antibody production. Our data extend those findings, demonstrating a role for these Tph cells in acute viral infections. We show that not only PD-1^high/int^CXCR5^+^ cTfh cells, but also PD-1^high^CXCR5^−^ Tph cells are significantly increased in peripheral blood of COVID-19 patients supporting the expansion of plasmablasts. One of the common signals between the acute phase of COVID-19 and chronic autoimmune conditions such as SLE are type-1 interferon (IFN) signatures^5,35,36^. Type-1 IFN (IFN-I) is reported to downregulate CXCR5 expression on human T cells^37^ and our recent data showed that IFN-I upregulates PD-1 expression and downregulates CXCR5 expression in an *in vitro* assay^38^. Intriguingly, most of the genes unique to PD-1^high^CXCR5^−^ Tph cells such as *CXCR6, LAG3, ZBTB20*, and *CHD7* had the same directions for their expression levels after type-1 IFN stimulation^38^. These data indicate that type-1 IFN might be one of the key signals to induce these Tph cells.

Patients with COVID-19 have a striking loss of GCs in lymph nodes with underlying extrafollicular B cell responses, particularly in severe cases^18,39^. Our scRNA-seq data showed an increase in FCRL5^+^ B cells in progressive patients (Supplemental Figure 1e), which supports this observation. Our results highlight the characteristics of PD-1^high^CXCR5^−^ Tph cells as: 1) lacking CXCR5 expression, important for the entry in GCs; 2) producing more IFNγ that is similar to T cells residing in the extra-follicular regions of lymph nodes^18^; and 3) exhibiting highly overlapping signatures with dividing CD4^+^ T cells that are positively correlated with the proportion of Ki67^+^ plasmablasts^40^, which had lower SHM frequencies in our scRNA-seq data. These findings suggest that circulating PD-1^high^CXCR5^−^ Tph cells may be the counterpart of T cells which promote more extra-follicular responses^20^. On the other hand, we could detect these T cells in peripheral blood of stable COVID-19 patients despite the lack of extrafollicular signatures in such patients. In acute viral infections, extra-follicular responses are thought to bridge between innate- and GC-responses of B cells^41^. Many effector molecules control the switch between GC- and extra-follicular responses^42^, which is dynamically changed in the acute phase of COVID-19. Though it might be difficult to conclude the direct relationship between extra-follicular responses and blood PD-1^high^CXCR5^−^ Tph cells from our dataset, the longitudinal assessment of these T cells together with their spatial analysis with B cells in lymph nodes will clarify these relationships.

In the *Salmonella* infected mouse model, the antibody responses important for the bacteria clearance are established by day 3 through 5 weeks, and GC formation is observed approximately one month after infection, indicating GC formation is not necessary for these early responses^43^. Similarly, in COVID-19 human samples, earlier antibody responses are likely to be associated with better recovery^44,45^, which is consistent with our findings that the rapid induction of inflammatory tissue-homing plasmablasts was linked with a better clinical outcome. Additionally, we detected that PD-1^high^CXCR5^−^ Tph cells in the activated state had the capacity to support the induction of tissue-homing plasmablasts, and these T cells were significantly increased in stable patients at the earlier stage of disease. The significant difference in the frequency of activated PD-1^high^CXCR5^−^ Tph cells between stable and progressive patients disappears over time, indicating that the prompt induction of activated PD-1^high^CXCR5^−^ Tph cells might be critical for the promotion of tissue homing plasmablasts to effectively remove virus from inflamed tissues and thus linked with better clinical outcomes. Not only sex differences^3^, but also genetic backgrounds are reported to be related to T cell activation^46,47^ and further analyses combined with these factors will lead to the more precise identification of patients at higher risk, and thus expected to be valuable for the development of personalized treatments.

Finally, our investigations have implications for understanding T-B cell interaction in human viral infections and provide a potential framework for assessing immune response to SARS-CoV-2 and other IFN-I inducing viral infections. In summary, our data implicate PD-1^high^CXCR5^−^ Tph cells as triggering acute protective plasmablast responses with direct relevance in COVID-19 pathophysiology. Moreover, these data shed light on how T cells can affect B cell differentiation in COVID-19 and provide potential insights into the role of PD-1^high^CXCR5^−^ Tph cells on a variety of immune-mediated diseases with possible contribution of aberrant T-B interaction, including chronic autoimmune diseases.

## Methods

### Ethics Statement

This study was approved by the Institutional Review Board at the Yale School of Medicine (2000027291REG and FWA00002571, Protocol ID. 2000027690). Informed consent was obtained from all enrolled patients, healthcare workers and healthy donors.

### Patients and Samples

Adult patients (≥18 years old) admitted to Yale-New Haven Hospital, positive for SARS-CoV-2 by RT-PCR from nasopharyngeal and/or oropharyngeal swabs, and able to provide informed consent (surrogate consent accepted) were eligible (Supplemental Table 1). Individuals with active chemotherapy against cancers, pregnant patients, patients with background hematological abnormalities, patients with autoimmune diseases and patients with a history of organ transplantation and on immunosuppressive agents, were excluded from this study. For the characterization of T cells and B cells, the flow data deposited in the Yale IMPACT Biorepository study were analyzed as described elsewhere^3,21^. All the patients were admitted between 30 March and 27 May 2020 and hospitalized. Only the baseline data were analyzed except for the analysis of time kinetics (Supplemental Table 2). All the experiments were performed on fresh peripheral blood mononuclear cells (PBMCs), and samples were drawn on the average of 11.8 days after first symptoms. 92 patients with COVID-19 and 64 COVID-19-uninfected healthcare workers (HCs) were enrolled. COVID-19 patients who required admission to the ICU had been classified as “progressive” and 27.8% of them were expired. The other patients classified as “stable” were all discharged without ICU admission. For the patients who are 90 years-old or older, their ages were protected health information, and ‘90’ was put as the surrogate value for the analyses. HCs were all negative in both PCR and serology tests.

Single cell RNA-seq (scRNA-seq) was performed on cryopreserved PBMC samples of 10 patients with COVID-19 following the same criteria as above and 13 age- and sex-matched controls. All the samples were drawn on the average of 11.7 days after first symptoms. Control samples were already collected before the first report of COVID-19 in 2018. From eight of ten patients with COVID-19, PBMC samples from two different time points had been analyzed. Four patients had been classified as “progressive”, who required admission to the ICU, and the other six patients classified as “stable” who were hospitalized and all discharged, and the same criteria as flow data. We have described the full cohort elsewhere^5^.

All the other experiments which include *in vitro* experiments and bulk RNA-seq were performed with fresh PBMCs at the baseline. All the patients were admitted between 21^st^ July and 22^nd^ Jan 2021.

### Peripheral blood mononuclear cells isolation

PBMCs were prepared from whole blood by Ficoll gradient centrifugation (Histopaque (Sigma) in the Yale IMPACT Biorepository study and Lymphoprep (Stemcell) in other experiments). The PBMC layer was collected into a new 50-ml tube and washed twice with PBS to remove any remaining Lymphoprep/Histopaque. As for scRNA-seq and flow cytometry, the pelleted cells were treated with ACK buffer for red cell lysis. All the other experiments including bulk RNA-seq samples were processed without lysis buffer.

### Flow Cytometry and sorting

In the Yale IMPACT Biorepository study, the staining was performed mainly in two separate panels for (1) T cell surface staining and (2) B cell surface staining. PBMCs were plated at 1-2 × 10^6^ cells in a 96-well U-bottom plate, and resuspended in Live/Dead Fixable Aqua (ThermoFisher) for 20 min at 4 °C. Following a wash, cells were then blocked with Human TruStan FcX (BioLegend) for 10 min at room temperature. Cocktails of desired staining antibodies were directly added to this mixture for 30 min at room temperature. Before analysis, cells were washed and resuspended in 100 μl of 4% paraformaldehyde for 30 min at 4 °C. We have described the detailed methods elsewhere^3^. The adopted antibodies and their clones were as follows. Anti-CD3 (UCHT1), anti-CD19 (SJ25C1), anti-CD4 (SK3), anti-CD45RA (HI100), anti-PD-1 (EH12.2H7), anti-CD38 (HIT2), anti-CXCR5 (RF8B2), anti-CXCR3 (1C6/CXCR3), anti-CD20 (2H7), anti-CD27 (M-T271), anti-IgD (IA6-2).

For other experiments, freshly isolated PBMCs were stained with cocktails of desired staining antibodies for 30 minutes at 4°C. Specific T cell and B cell subsets were sorted on a Sony MA900 cell sorter. The adopted antibodies and their clones were as follows. Anti-CD4 (OKT4), anti-CD19 (HIB19), anti-CD20 (2H7), anti-CD27 (M-T271), anti-CD38 (HIT2), anti-CD138 (MI15), anti-PD-1 (EH12.2H7), anti-LAG3 (11C3C65), anti-CCR2 (K036C2), ant-CCR5 (J418F1), anti-CXCR3 (G025H7), anti-CXCR6 (K041E5), anti-CX3CR1 (2A9-1) (all from Biolegend); anti-CD3 (UCHT1), anti-CD45RA (HI100), anti-CXCR5 (RF8B2) (all from BD Biosciences).

### T-B-cell co-culture experiments

Co-culture experiments were performed referenced as described previously^19^. In brief, sorted T cell populations (5000-10000 cells) from PBMCs of patients with COVID-19 were co-cultured with autologous CD20+CD27+ memory B cells at a ratio of 1:3 in 200 μl of RPMI 1640 medium (Gibco) supplemented with 10% fetal bovine serum (FBS), 2 nM L-glutamine, and 100 U/ml penicillin, 100 μg/ml streptomycin (Lonza), stimulated with SEB (1 μg/ml) and LPS (1 μg/ml) for 7 days. Supernatants were collected, and total IgG (Invitrogen) was measured by ELISA. Cells were harvested and analyzed by flow cytometry, with plasma cells defined as CD27^high^CD138^+^ cells.

### B cell differentiation *in vitro*

We extracted PBMCs from healthy volunteers. After the isolation of CD19+ B cells from PBMCs using Human B cell isolation kit (Stemcell Technologies), CD20+CD27+ memory B cells were sorted on a FACS Aria (BD Biosciences) and stimulated with CD40L (0.05 μg/ml) (Enzo), IL-21 (20 ng/ml) (R&D systems) and other cytokines (all from R&D systems) in culture medium the same as above. After 7 days, CD27^high^CD138+ plasma cells were sorted for gene expression analysis by qPCR.

### T cell stimulation *in vitro*

We extracted PBMCs from patients with COVID-19. Each subset of memory CD4+ T cells based on the expression levels of PD-1 and CXCR5 were stimulated with anti-CD3/CD28 (each 1 μg/ml; BD Biosciences) for 48 hrs and cytokine levels were measured by ELISA (all from R&D systems) according to the manufacturer’s instructions. 1% Triton X-100 for 60 minutes at room temperature was added before ELISA to reduce risk from any potential virus in the supernatant^48^.

### Quantitative PCR

Total RNA was extracted using RNeasy Micro Kit (QIAGEN) according to the manufacturer’s instructions. cDNA was synthesized with SuperScript IV VILO Master Mix (Invitrogen). cDNAs were amplified with Taqman probes (Taqman Gene Expression Arrays) and TaqMan Fast Advanced Master Mix on a StepOne Real-Time PCR System (Applied Biosystems) according to the manufacturer’s instructions. The RNA expression was measured relative to *B2M* expression. The adopted Taqman probes were as follows. *CXCR3* (Hs01847760_s1), *CXCR4* (Hs00607978_s1), *CCR2* (Hs00356601_m1).

### Single cell RNA-seq data processing

A PBMC scRNA-seq data set which had been previously performed and reported by us^5^ was reanalyzed. In brief, single cell barcoding of PBMC and library construction had been performed using the 10x Chromium NextGEM 5 prime kit according to manufacturer’s instructions. Libraries had been sequenced on an Illumina Novaseq 6000 platform. Raw reads had been demultiplexed and processed using Cell Ranger (v3.1) mapping to the GRCh38 (Ensembl 93) reference genome. Resulting gene-cell matrices had been analyzed using the package Seurat^49,50^ in the software R (v3.6.2) including integration of data, clustering, multiplet identification and cell type annotation. We have described the detailed methods elsewhere^5^. The annotated R object was used for sub-clustering of B cells.

The three cell populations, “Memory B cells”, “Naïve B cells “ and “Plasma cells” in total PBMCs were re-clustered to obtain a finer cell type granularity. To remove batch- and single-donor effects, we integrated all 31 samples of these populations into one dataset using reference-based anchor finding and integration workflow. We chose 2 healthy donor samples (C27 and C32) and 2 COVID-19 samples (NS1B and TS3A), which have enough B cell numbers, as references for anchor finding and integration. The top 2000 variable genes were selected, and integration anchors were determined by “FindIntegrationAnchors” without k.filter for low cell numbers in some samples. These anchors were used to integrate the data using the “IntegrateData” function with top 30 dimensions and scaled. The top 17 PCs were used for data integration and downstream steps, along with a clustering resolution of 0.4. Cluster-specific gene expression profiles were established using the “FindAllMarkers” per cluster and per subset to annotate the clusters. Doublet clusters were determined by co-expression of heterogeneous lineage markers (e.g., *MS4A1* and *CD3*). These clusters were removed prior to finalizing the UMAPs.

### B cell receptor repertoire analysis

B cell receptor (BCR) V(D)J repertoire data processing, clonal clustering, and unmutated germline ancestor sequence reconstruction was previously performed in^5^. Briefly, V(D)J genes aligned to the IMGT/GENE-DB v3.1.26^51^ germline reference database using IgBLAST v.1.15.0^52^. Cells with multiple IGH V(D)J sequences were assigned to the most abundant IGH V(D)J sequence by UMI count, and ties were broken by the first identified heavy chain. Non-functional sequences were removed. V(D)J sequences within each patient were grouped into clonal clusters by first partitioning based on common IGHV gene annotations, IGHJ gene annotations, and junction lengths. Within these groups, sequences differing from one another by a length normalized Hamming distance of 0.15 within the junction region were defined as clones by single-linkage clustering using Change-O v.1.0.0^53^. Germline sequences were reconstructed for each clone with the D segment and N/P regions masked (replaced with “N” nucleotides) using the CreateGermlines.py function within Change-O v.1.0.0.

Somatic hypermutation frequency was calculated using SHazaM v1.0.2.999^54^ as the frequency of non-ambiguous nucleotide differences along the IGHV gene segment (IMGT positions 1-312) between each sequence and its inferred germline ancestor. To identify unmutated B cell clones of different cell types and isotypes, B cell clones were separated by cell type and isotype and considered “unmutated” if the median somatic hypermutation frequency of their constituent sequences was < 1%. This cutoff was also used in^5,55^. To quantify B cell clonal diversity, we calculated Simpson’s diversity within each patient for plasmablast subsets using the alphaDiversity function of Alakazam v1.0.2.999^53^. To account for differences in sequence depth, the number of sequences within each patient were down-sampled to the same number of sequences, and the mean of 100 such re-sampling repetitions was reported. Only patients with at least 30 B cells were included in diversity calculations. All statistical analyses of BCR sequences were performed with R (v3.6.1).

### Bulk RNA-seq

#### cDNA and library preparation and sequencing

RNAs were isolated using RNeasy Plus Micro Kit (QIAGEN) and cDNAs were generated using the SMART-Seq v4 Ultra Low Input RNA Kit for sequencing (Takara/Clontech).

Barcoded libraries were generated by the Nextera XT DNA Library Preparation kit (Illumina) and sequenced with a 2×100 bp paired-end protocol on the HiSeq 4000 Sequencing System (Illumina).

#### Bulk RNA-seq data analysis

Low quality ends (less than phred score=30) and short read length (minimum length=30) was trimmed using PRINSEQ++^56^ (version1.2). Trimmed reads were aligned to the hg38 genome reference using STAR^57^ (v2.7.1), and subsequently RSEM (RNA-Seq by Expectation-Maximization)^58^ was used to count reads mapping to the genes from Ensembl release 93. Top 1000 genes by variance were analyzed for PCA. Heat maps show row-normalized relative gene expression z-scores across columns. Pairwise differential expression was performed using the R package DESeq2^59^. The cutoff value to select differentially expressed genes is provided in each figure legend.

These data will be publicly available prior to publication.

## Statistical analysis

All statistical analyses were performed using R or GraphPad Prism 7 (GraphPad Software). Detailed information about statistical analysis, including tests and values used, is provided in the figure legends.

## Data Availability

The scRNA-seq and bulk RNA-seq data will be available before publication in a public repository.
Flow cytometry data are available via ImmPort website (study ID; SDY1648, SDY1655). Data used to generate Figures are included in Supplemental Tables.

## Supplemental Figure Legends

**Supplemental Figure 1.**
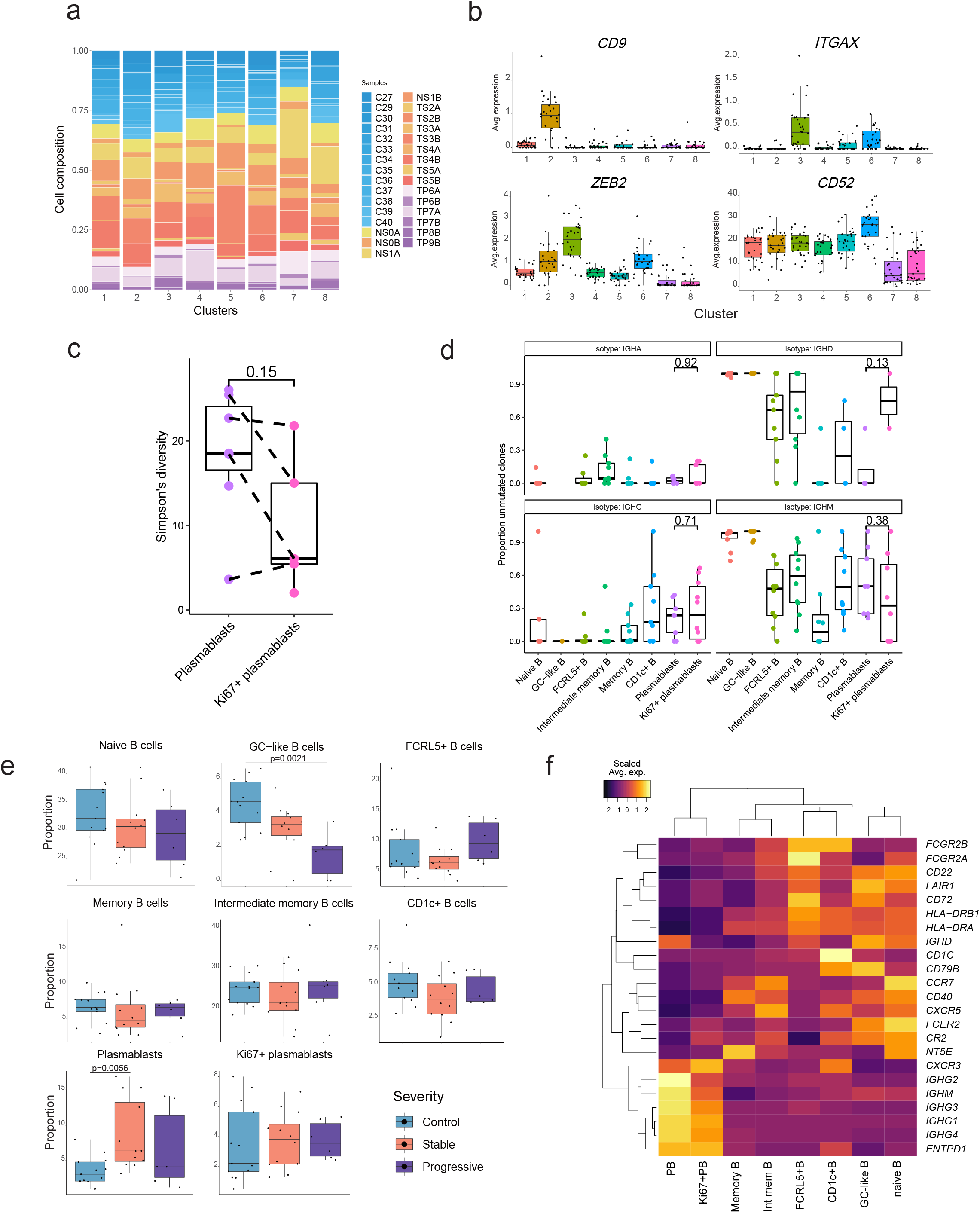
The characteristics of each B cell subset in scRNA-seq dataset. **a**, Bar plot showing cell compositions of each cluster by samples. **b**, Box plot showing other canonical markers. The median is marked by a horizontal line with whiskers extending to the farthest point within a maximum of 1.5 x interquartile range. Each dot corresponds to each sample. **c**, Simpson’ s diversity of B cell clones within each plasmablast cluster. Each dot corresponds to a patient (combined early and late samples), and dots from the same patient are connected with dotted lines. **d**, The proportion of unmutated clones within each cell type cluster based on immunoglobulin isotypes. Each dot corresponds to a patient, and a Wilcoxon test p value is reported above plasmablast clusters. **e**, Comparison of cell counts (percentage of total B cells) among each group (one-way ANOVA with Dunnet’ s multiple comparisons test). Each dot corresponds to each sample, and One-way ANOVA with Dunn’ s multiple comparisons test was performed. **f**, Heatmap of gene expressions related to B cell functions^31^ in each cluster. All the samples are evaluated.

**Supplemental Figure 2.**
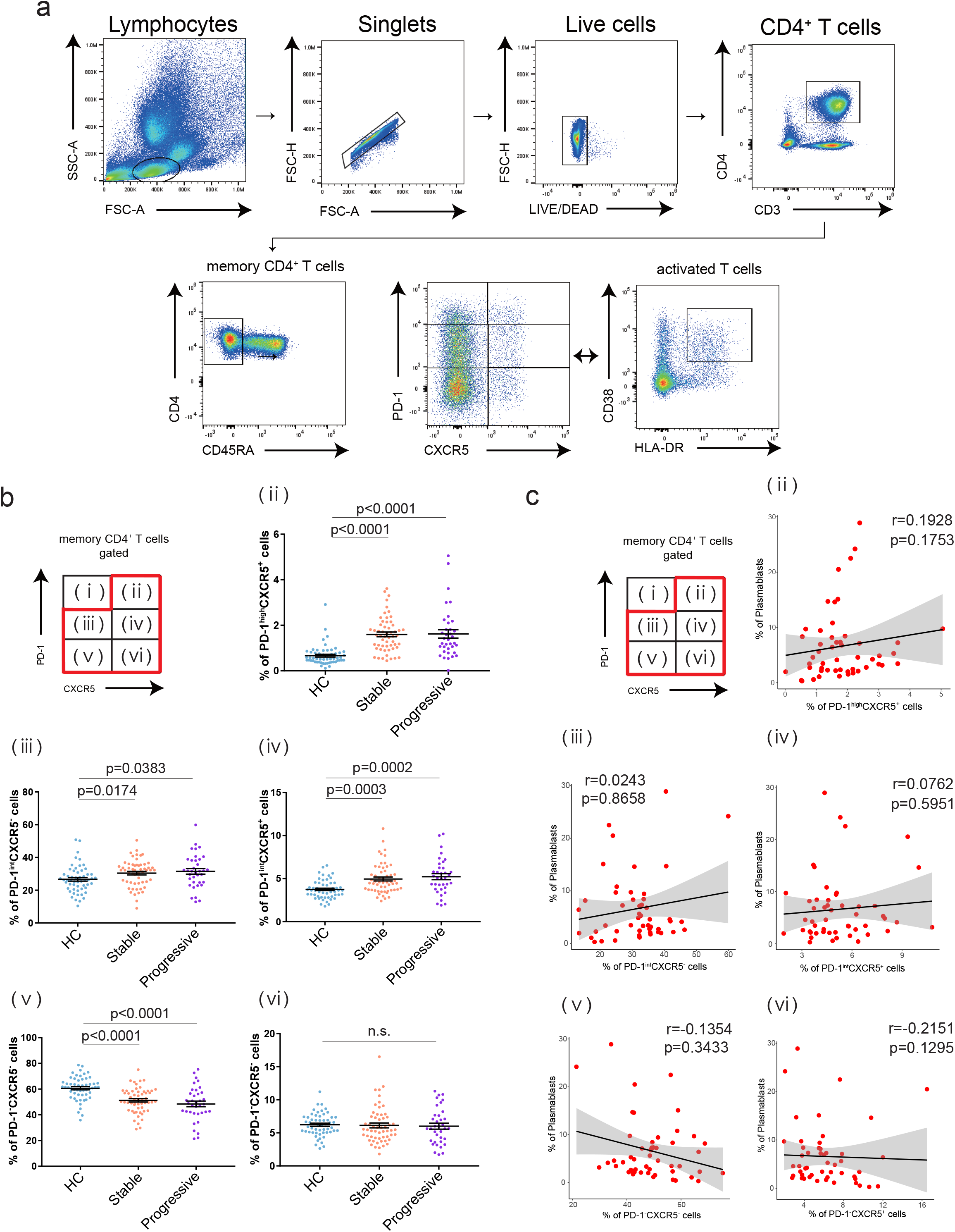
The characteristics of each T cell subset. **a**, Gating strategy to identify each T cell subset in PBMCs. Six subsets were detected based on the expression levels of PD-1 and CXCR5, and activated T cells were defined as HLA-DR^+^CD38^+^ T cells. **b**, The proportion of each T cell subset among healthcare workers (HC) (n=55), stable COVID-19 patients (Stable) (n=56), and progressive patients (Progressive) (n=36). One-way ANOVA with Dunn’ s multiple comparisons tests were evaluated. n.s. = no significance among each group. **c**, Correlation between each T cell subset (percentage of CD3^+^CD4^+^CD45RA-memory T cells) and plasmablasts (percentage of CD19^+^ B cells) in COVID-19 patients (both stable and progressive, n=51). Linear regression is shown with 95% confidence interval (gray area). Correlation statistics is two-tailed Spearman’ s rank correlation test.

**Supplemental Figure 3.**
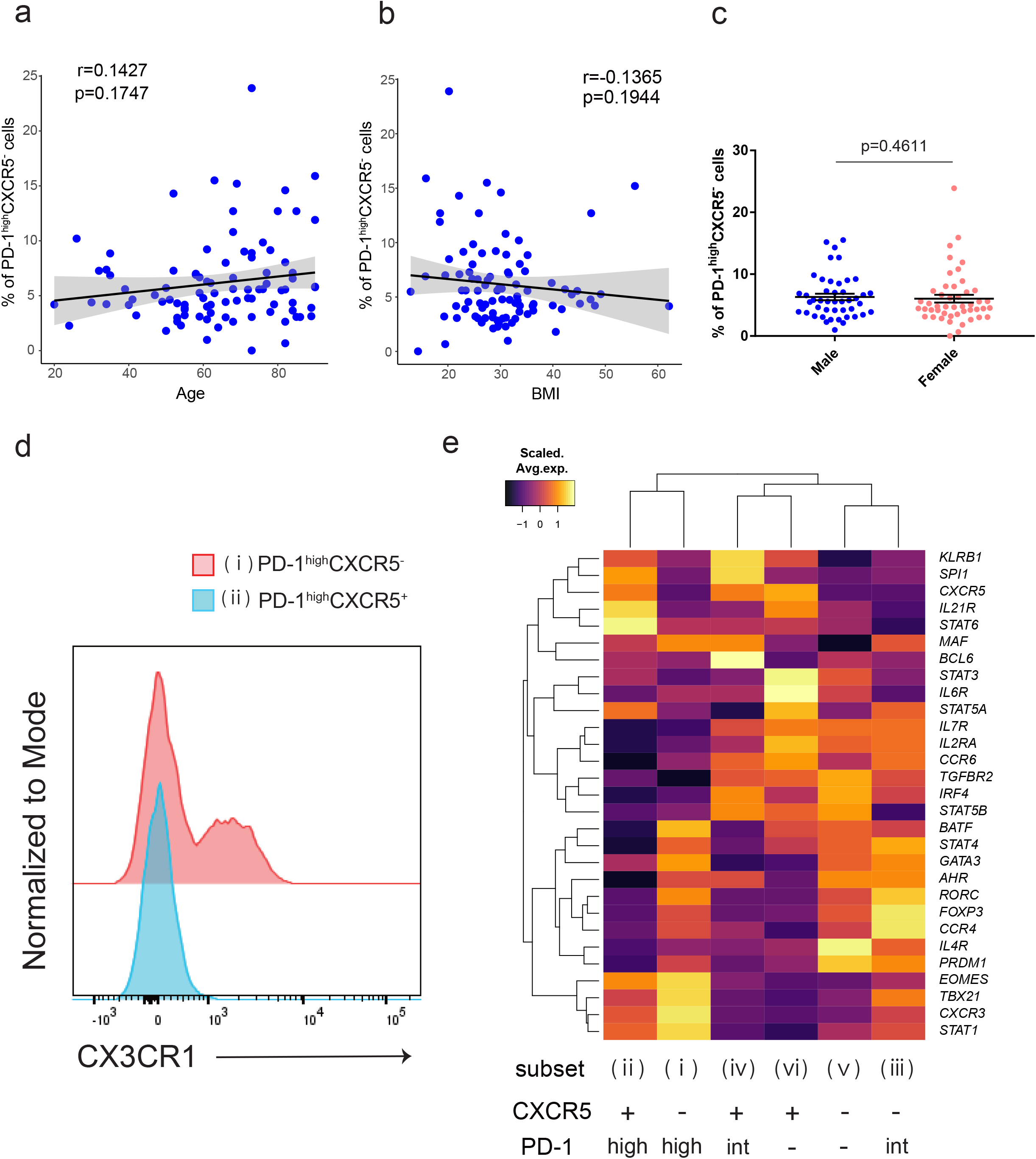
The characteristics of PD-1^high^CXCR5^−^ Tph cells. **a-b**, Correlation between the proportion of PD-1^high^CXCR5^−^ Tph cells (percentage of CD3^+^CD4^+^CD45RA-memory T cells) and each clinical background in COVID-19 patients (both stable and progressive, n=92) (**a**, age; **b**, BMI). Linear regression is shown with 95% confidence interval (gray area). Correlation statistics by two-tailed Spearman’ s rank correlation test (**a, b**). **c**, The proportion of PD-1^high^CXCR5^−^ Tph cells between male (n=45) and female (n=47) COVID-19 patients were evaluated by two-tailed unpaired Student’ s t-test. **d**, Representative flow data of CX3CR1 expression on PD-1^high^CXCR5^−^ Tph cells compared with PD-1^high^CXCR5^+^ Tfh cells. **e**, Heatmap of T cell lineage genes among six subsets of memory CD4^+^ T cells.

**Supplemental Figure 4.**
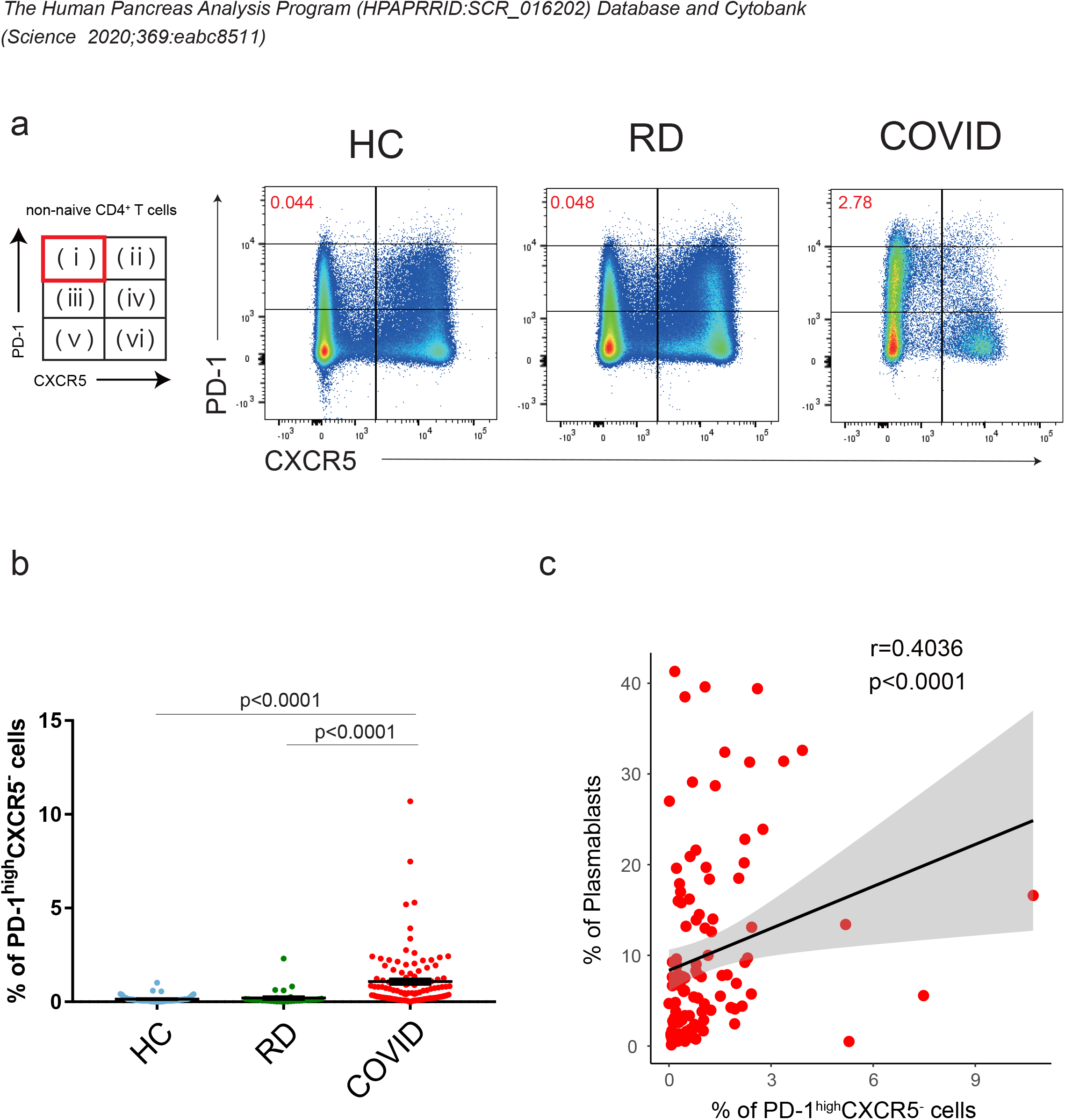
PD-1^high^CXCR5^−^ Tph cells from another COVID-19 dataset. Deposited flow cytometry data from another study^6^ were analyzed for validation of the characteristics of PD-1^high^CXCR5^−^ Tph cells in the acute phase of COVID-19 patients. **a**, Representative flow data of PD-1^high^CXCR5^−^ Tph cells in healthy donors (HC), recovered donors from COVID-19 (RD), and hospitalized COVID-19 patients (COVID). **b**, The proportion of PD-1^high^CXCR5^−^ Tph cells among HC (n=56), RD (n=36), and COVID (n=109; all at baseline samples) groups. One-way ANOVA with Dunn’ s multiple comparisons tests were performed to evaluate differences. **c**, Correlation between PD-1^high^CXCR5^−^ Tph cells (percentage of CD3^+^CD4^+^CD45RA^−^ non-naive T cells) and plasmablasts (percentage of CD19^+^ B cells) in COVID-19 patients (n=109). Linear regression is shown with 95% confidence interval (gray area). Correlation statistics is two-tailed Spearman’ s rank correlation test.

**Supplemental Figure 5.**
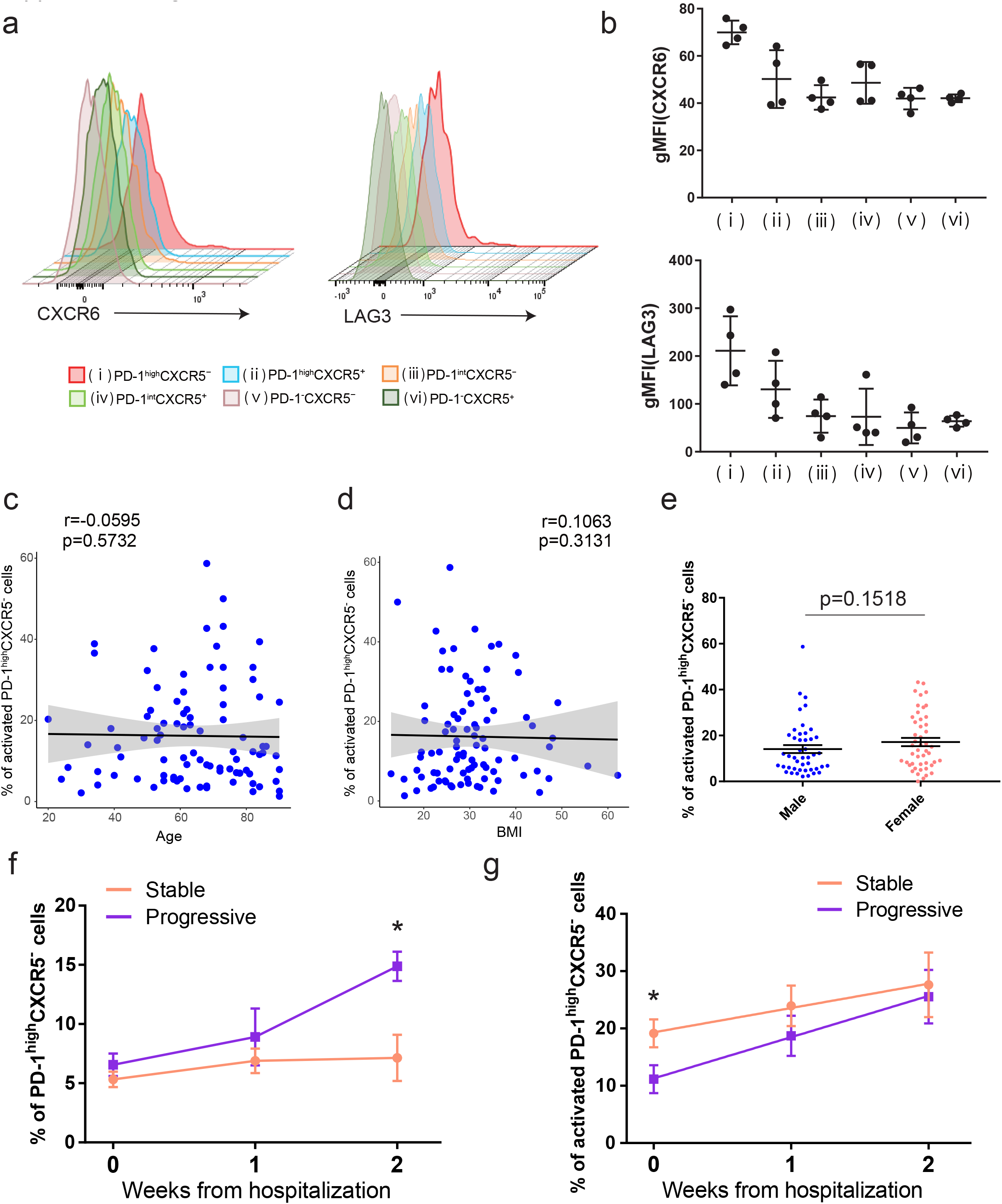
The characteristics of HLA-DR^+^CD38^+^ activated PD-1^high^CXCR5^−^ Tph cells. **a**, Representative flow data of CXCR6 and LAG3 on each T cell subset. **b**, LAG3 (left) and CXCR6 (right) gMFI of each T cell subset were evaluated (n=4, COVID-19 patients). **c-d**, Correlation between the proportion of activated HLA-DR^+^CD38^+^PD-1^high^CXCR5^−^ Tph cells (percentage of PD-1^high^CXCR5^−^ Tph cells) and each clinical background in COVID-19 patients (both stable and progressive, n=92)(**c**, age; **d**, BMI). Linear regression is shown with 95% confidence interval (gray area). Correlation statistics is two-tailed Spearman’ s rank correlation test (**c, d**). **e**, The proportion of activated HLA-DR^+^CD38^+^PD-1^high^CXCR5^−^ Tph cells between male (n=45) and female (n=47) COVID-19 patients were evaluated by two-tailed unpaired Student’ s t-test. **f-g**, Longitudinal frequencies of PD-1^high^CXCR5^−^ Tph cells (**f**) and activated PD-1^high^CXCR5^−^ Tph cells (**g**) after hospitalization. Only the samples which could follow blood collection (hospitalization, week1 of day1-7, week2 of day8-14) were analyzed (Stable n=23, Progressive n=16). At each time point, Two-tailed unpaired Student’ s t-test were performed (*p<0.05).

**Supplemental Figure 6.**
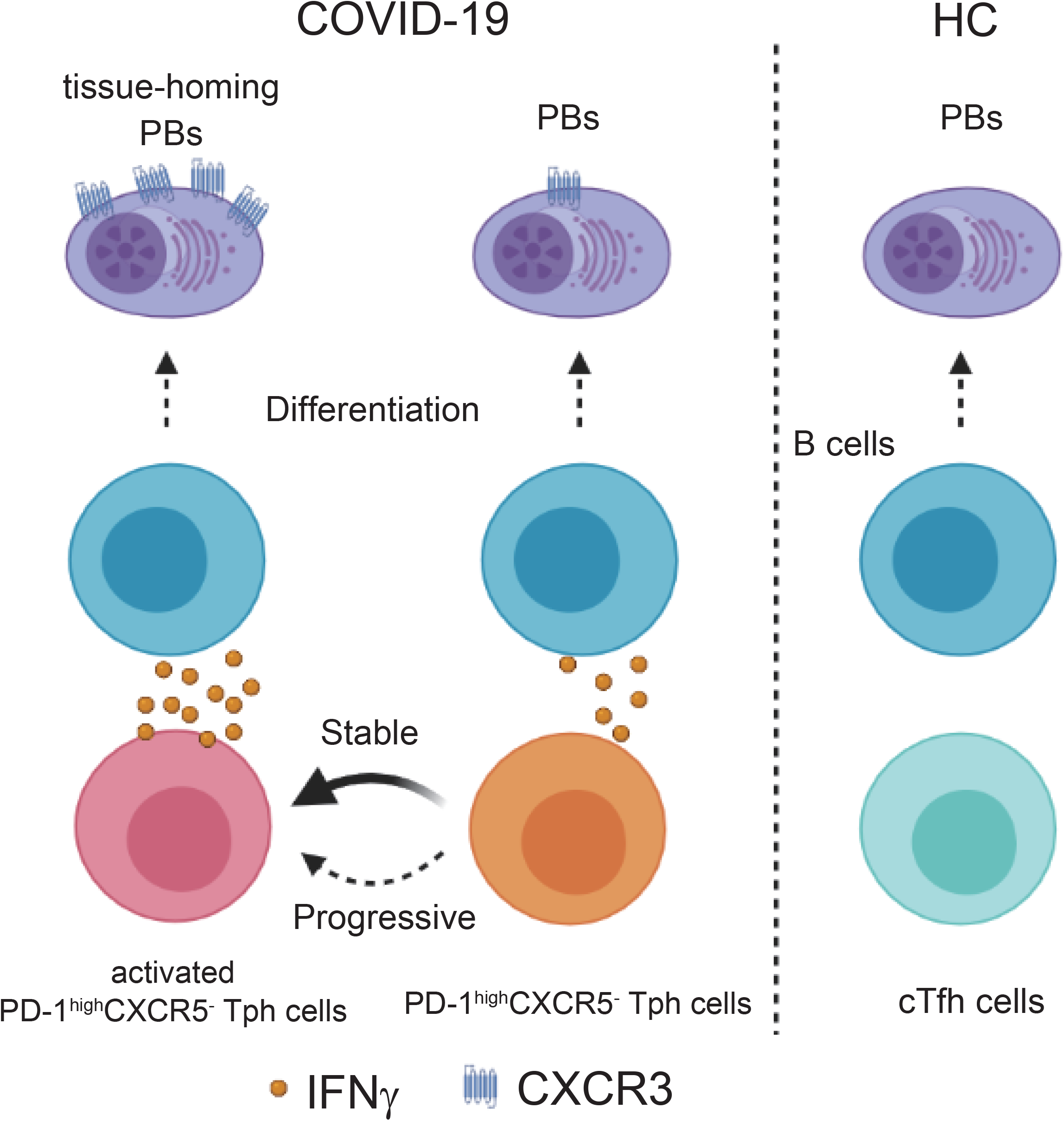
Schematic model of T-B interactions in a) healthy donors, b) stable COVID-19 patients, and c) progressive COVID-19 patients in acute phase. Under healthy conditions, we can detect few plasmablasts and PD-1^high^CXCR5^−^ Tph cells. In the acute phase, PD-1^high^CXCR5^−^ Tph cells increased in COVID-19 patients and are related to the promotion of plasmablasts. Among patients with COVID-19, stable groups can increase activated PD-1^high^CXCR5^−^ Tph cells quickly, which can produce IFNγ more than PD-1^high^CXCR5^+^ Tfh cells, and promote inflammatory-tissue homing plasmablasts at the proper timing. Progressive COVID-19 patients can also increase activated PD-1^high^CXCR5^−^ Tph cells but delayed, which lead to insufficient promotion of plasmablasts at proper timing and worse clinical outcome.

## Acknowledgments

We would like to thank all the hospital staff who helped care for the patients and obtain samples. We are also grateful to all the members of YALE IMPACT research team who obtained data. We also thank Yale Environmental Health and Safety (EHS) office, particularly Dr. Maren Schniederberend, for providing the safety guidance for working with COVID19 samples; Dr. Kevin O’Connor and C. Philip for feedback and discussions; Drs. L. Devine and C. Wang for assistance with FACS based cell sorting; Drs. G. Wang and C. Castaldi at Yale Center for Genome Analysis for support with 10x Genomics library preparation and sequencing. We also thank Mei Zhang for preparation of bulk RNA-seq libraries and sequencing. H.A. thanks Daiichi Sankyo Foundation of Life Science and Uehara Memorial Foundation for his scholarship. D.A.H, A.I., S.H.K., R.R.M., and A.C.S. thank the HIPC Consortium for valuable input.

## Competing interest statement

D.A.H. has received research funding from Bristol-Myers Squibb, Novartis, Sanofi, and Genentech. He has been a consultant for Bayer Pharmaceuticals, Bristol Myers Squibb, Compass Therapeutics, EMD Serono, Genentech, Juno therapeutics, Novartis Pharmaceuticals, Proclara Biosciences, Sage Therapeutics, and Sanofi Genzyme. Further information regarding funding is available on:

https://openpaymentsdata.cms.gov/physician/166753/general-payments

N.K. reports personal fees from Boehringer Ingelheim, Third Rock, Pliant, Samumed, NuMedii, Indalo, Theravance, LifeMax, Three Lake Partners, RohBar in the last 36 months, and Equity in Pliant. N.K. is also a recipient of a grant from Veracyte and nonfinancial support from Miragen. In addition, N.K. has patents on New Therapies in Pulmonary Fibrosis and ARDS (unlicensed) and Peripheral Blood Gene Expression as biomarkers in IPF (licensed to biotech) all outside the submitted work. S.H.K. receives consulting fees from Northrop Grumman.

## Author Contributions

Overall study design; H.A., D.A.H., T.S.S.

Biospecimen collection/processing; H.A., M.S., M.C., W.E.R., P.W., K.R., O.C., A.U., B.E.,

R.R.M., A.I., A.C.S, T.S.S.

Data analysis; H.A., W.R., K.B.H., I.C., S.C., S.H.K., C.D.C., N.K.,

Original draft writing; H.A.

Supervising the study; S.H.K., C.D.C., N.K., A.C.S, D.A.H., T.S.S.

Reviewing and editing the manuscript; All authors participated in editing the manuscript.

